# Frequency and accuracy of proactive testing for COVID-19

**DOI:** 10.1101/2020.09.05.20188839

**Authors:** Ted Bergstrom, Carl T. Bergstrom, Haoran Li

## Abstract

September 5, 2020

The SARS-CoV-2 coronavirus has proven difficult to control not only because of its high transmissibility, but because those who are infected readily spread the virus before symptoms appear, and because some infected individuals, though contagious, never exhibit symptoms. Proactive testing of asymptomatic individuals is therefore a powerful, and probably necessary, tool for preventing widespread infection in many settings. This paper explores the effectiveness of alternative testing regimes, in which the frequency, the accuracy, and the delay between testing and results determine the time path of infection. For a simple model of disease transmission, we present analytic formulas that determine the effect of testing on the expected number of days of during which an infectious individual is exposed to the population at large. This allows us to estimate the frequency of testing that would be required to prevent uncontrolled outbreaks, and to explore the trade-offs between frequency, accuracy, and delay in achieving this objective. We conclude by discussing applications to outbreak control on college and university campuses.

**Competing Interest Statement:** Ted Bergstrom and Haoran Li have no competing interests. Carl Bergstrom consults for Color Genomics on COVID testing schedules.

---

The SARS-CoV-2 coronavirus has infected six million people in the United States as of late August 2020. Unlike the SARS virus which appeared in 2003 [1, 2], SARS-CoV-2 is readily transmitted before symptoms develop [3, 4, 5]. Entirely asymptomatic infections are common [6, 7, 8], yet asymptomatic patients transmit disease. [9, 2, 10, 11]. The severity of COVID varies from mild illness [12] to pneumonia and acute respiratory distress syndrome [13]. Symptoms are highly variable, and range from respiratory to gastrointestinal, neurological to circulatory [14, 15]. As a consequence, neither symptomatic screening, nor self-isolation based on symptoms is likely to identify the majority of cases [16, 17]. As a result, frequent proactive testing will be an important component of control, because it can provide a surrogate for symptomology in identifying and subsequently isolating infectious individuals [18].

To the first order, the early dynamics of an epidemic are shaped by a single parameter, the “basic reproduction number” *R*_0_. This quantity is defined as the expected number of secondary infections arising from an index case in a wholly susceptible population [19]. In the initial stages of an epidemic the number of cases can be expected grow exponentially over time when *R*_0_ > 1. When *R*_0_ < 1, newly introduced infections will result in only a limited number of cases. As an epidemic progresses and the number of susceptible individuals declines, we look at the effective reproductive number *R* = *R*_0_ *S*, where *S* is the fraction of the population that remains susceptible to disease. To control the spread of disease, a community, workplace, university, or other setting needs to ensure that *R* remains below unity.

The basic reproduction number is not an intrinsic property of the virus, but rather reflects the organization and social behavior of a given population at a particular time. Thus the basic reproduction number varies across subpopulations, and changes in response to infection control measures. A commonly used estimate of *R*_0_ for COVID, in the absence of social distancing measures, is *R*_0_ = 2.5 [20]. Non-pharmaceutical interventions such as face masks, social distancing, and limits on large gatherings can reduce *R*_0_ somewhat, but are unlikely to drive the basic reproduction number below unity without taking a dramatic toll on the social and economic life of a community. Frequent proactive testing of asymptomatic individuals offers an additional way to reduce transmission, and is among the most powerful and least disruptive of non-pharmaceutical interventions.

How much testing is necessary to reduce *R* by a given amount? This depends in intricate ways on the biology of the virus and the details of the population in question—but a simple heuristic provides a nice approximation. Imagine that each infectious person is either at large in the community or isolated at home during each day of the infectious period. The number of contacts that an infectious person has will be roughly proportional to the fraction of the infectious period spent at large. If transmission is eliminated entirely while isolated, then to reduce *R* to 1, it is necessary that reduce the average fraction of the infectious period spent at large to 1*/R*. This means that if *R* = 2.5 in the absence of testing, then to achieve *R* < 1 through testing and isolation, it would be necessary to reduce the fraction of the time that infectious individuals are at large in the community by more than 60 percent.

Communities face considerable variation and uncertainty regarding COVID transmission rates, and in the effects that non-pharmaceutical interventions have on transmission. There is also substantial variation (and some uncertainty) in the sensitivities and specificities of alternative testing methods, in the costs of these methods, and in the turnaround times that these methods offer between testing and reporting. The interactions between these effects is complex and non-linear. In this paper, we develop analytic formulas that allow one to calculate the predicted reduction in transmission rates that result, under alternative parameters of infection from any specified testing regime.

## Testing regime and contagious exposure

Testing plays multiple roles in pandemic control. Testing for individual diagnosis is used to determine whether patients exhibiting disease symptoms are suffering from COVID or some other malady. Point-of-care testing can serve to clear individuals to undergo medical or dental procedures, or engage in other high-contact activity. Testing serves a surveillance role, helping public health officials track the prevalence of disease and the rate of spread. Finally, by proactively testing individuals who are not showing symptoms, health workers can identify individuals who are infected but do not realize it, and isolate them from interactions with the community. We focus on this last role here.

Throughout our discussion, we will assume that the course of coronavirus infection can be parameterized as follows:

**Assumption 1** (Parameters of infection). *A vector of infection parameters γ* = (*C, u, v, y*) *is defined in the following way:*

- *The infectious period for all cases, symptomatic or asymptomatic, is C days*.
- *A fraction u of cases are asymptomatic*
- *Among symptomatic cases, a fraction v self-isolate once the symptoms appear and will no longer transmit disease*.
- *Among symptomatic cases, there is a pre-symptomatic period y days during which a patient is infectious. This includes the time from the onset of the symptoms until the time that the patient realizes that isolation is warranted*.

Because symptoms vary and mild COVID symptoms can be confused with seasonal allergies, a common cold, or other maladies, a majority of symptomatic individuals may not quarantine. Larremore et al [21] assume that only 20% of those who are infected self-isolate when symptoms appear; Color Genomics [22] uses a slightly higher value of 30% for a workplace population.

Infected individuals who become symptomatic are contagious for an average of approximately 3 days before symptoms appear [23]. There is substantial divergence in estimates of the average number of days of infectiousness after symptoms appear. The CDC^1^ cites studies that indicate that for those with mild symptoms, the number of days of infectiousness after infection is “less than 10” according to one source and “less than 6” according to another source. Adding the number of days of presymptomatic contagion to the post-symptomatic number, it is reasonable to suppose that, on average, an infected person is infectious for 7 or 8 days.

Some infected individuals remain fully asymptomatic for the entire course of infection, and never develop any noticeable symptoms. The CDC estimates that approximately 40% of infected persons fit into this category [20].

We have limited information about the sensitivity (defined as one minus the false negative rate) of RNA-based tests for COVID, and much of what we do have is drawn from patients with serious illness. Estimating test sensitivities for pre-symptomatic and asymptomatic persons is especially difficult; we have found no studies that directly address the sensitivity for these populations. However, contact tracing studies [2], [10] indicate that asymptomatic and pre-symptomatic individuals carry similar viral loads to those with symptoms and thus are comparably likely to transmit disease. A survey of studies based on Chinese hospital patients [24] reports test sensitivities ranging from 71% to 98%. A survey of research that includes some out-patient and in-patient cases by Kucirka *et al*. [25] estimates that the fraction of false-negatives ranges from 38% when symptoms first appear to 20% when symptoms disappear.^2^ In a brief article advising physicians on the interpretation of test results, Watson et al [26] observe that “As current studies show wide variation and are likely to overestimate sensitivity, we will use the lower end of current estimates from systematic reviews, with approximate sensitivity of 70%” (equivalently false-negative rate of 30%.)

Given the considerable uncertainty around the test sensitivities and the variation across sample collection and testing methods, we treat this rate as a parameter which can be adjusted when calculating the effects of testing.

We parameterize a testing regime as follows.

**Assumption 2** (Parameters of the testing regime). *A testing regime τ* = (*n, q, d*) *is characterized as follows:*

- *Everyone is tested at regular intervals, where n is the number of days between tests*.
- *An infectious individual will have a false negative test result with probability q. (I.e., test sensitivity is* 1 *− q.) The probability that an infectious person has a false negative result on any test is independent of the results of prior tests taken while infectious*.
- *After a delay of d days from the time of the test, those who test positive will be isolated for the remainder of their infectious period*.

**Definition 1** (Expected exposure function). *The expected exposure function E*(*C, τ*) *is the expected amount of time that someone will be contagious and at large under testing regime τ if, in the absence of testing, they would be contagious and at large for a period of C*.

The expected exposure time depends on the parameters of infection and of the testing regime in a rather complex, non-linear way. In subsequent discussion, we find explicit formulas for the expected exposure function. We have developed a “Contagion Calculator” which can be found at https://steveli.shinyapps.io/FrequencyAndAccuracyCalculator/. With this calculator, one can input values for the parameters of infection and of the testing regime, and the the calculator will output expected exposure days for an infected person, the ratio of expected exposure days with testing to that with no testing, and an estimated effective reproductive number *R* that would be achieved with this testing regime.

## Exposure ratios when testing intervals are longer than the period of infectiousness

If testing intervals are longer than the period of infectiousness, then some infected persons will never be tested and those who are tested will be tested at most once while contagious. This makes calculation of exposure ratios relatively easy, but also means that testing will have only a small effect on the expected number of days that infected persons are exposed to the public while contagious. For example, at least one major university plans to test asymptomatic persons just once per month upon reopening^3^ Our calculations show that with monthly testing, if all of those who test positive are isolated, will reduce their effective reproductive number *R* by less than 10 per cent.

If the interval *n* between tests is longer than *C − d*, an infected person will be tested at most one time while contagious. Let the random variable t be the length of the period between the time when a subject first becomes infectious and the time of the next test. We assume that t is uniformly distributed on the interval [0, *n*] between tests.^4^ The results of a test for someone who has t = *t > C −d* will become available only after this person is no longer infectious. Thus this person will be contagiously exposed for the full period *C* of contagion. If *t < C − d*, then the subject will be infected when tested and will test positive and be isolated with probability 1 *− q*. At the time of testing, the subject will have been contagiously exposed for a period of *t* before the test and will continue to be so for an additional time *d* until test results appear. Thus if the test reports accurately, the total time of contagious exposure will be *t* + *d*. It follows that the expected amount of time of contagious exposure for an infected person is

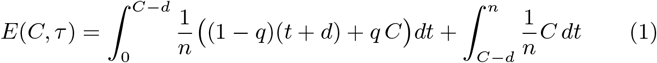

As we show in the Appendix, Equation 1 simplifies to yield the following result:

**Proposition 1.** *Given Assumption 2, if n > C − d*,

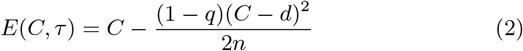

## Exposure ratios when testing intervals are shorter than the period of infectiousness

When testing intervals are shorter than the period of infectiousness, every infected person will be tested at least once during their infectious period. Some, who are infected but test negative, may be tested again in time for the results to be returned while they remain infectious. We assume that the probability of a negative result for an infectious person on any test is independent of the results of previous tests.

We model the the amount of time between the onset of infectiousness and the first time that a subject is tested as a random variable t drawn from a uniform distribution on the interval [0, *n*]. Suppose that the time interval from the beginning of infectiousness to a subject’s first test is *t < C − d*. With probability 1 *− q*, the test will correctly report a positive result. In this case, the subject will be isolated with a time delay of *d* and will have been contagiously exposed to the public for a total duration of *t* + *d*. With probability *q*, the first test incorrectly reports a negative result. In this case, the subject test will be tested again after *n* days and every *n* days thereafter so long as he or she continues to test negative. For a subject who is first tested at an interval of *t* after becoming infectious, let *x*(*t*) be the number of testing occasions that occur while this patient remains infectious and would still be infectious when the test results come back. This is the largest integer *i* such that *t* + *d* + *n i ≤ C*. Equivalently, this is the largest integer *i* such that *i ≤* (*C −t−d*)*/n*. We use a standard notation *b·c* for the “floor” function to write the following definition:

**Definition 2.** *Where* 0 *≤ t ≤ C − d, let 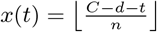. Let* 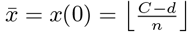, *and let* 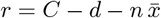.

When 0 *≤ i ≤ x*(*t*), the probability that someone whose first test occurs after being contagious for length of time *t*, who has *i* consecutive false negative results, and who tests positive on the next occasion, is (1 *− q*)*q^i^*. In this event, the number of days of infectious exposure would be *t*+*n i*+*d ≤ C*. With probability *q^x^*^(*t*)+1^, an infectious person will never be isolated while contagious, and thus will be infectious and at large for *C* days.

It follows that where the contagious period is *C* days and the testing regime is *τ* = (*n, q, d*), the expected number of days of contagious exposure for someone who is first tested after being contagious for a length of time *t* is

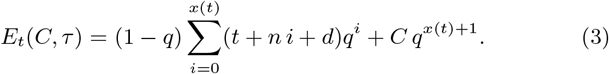

Since *t* is assumed to be uniformly distributed on the interval [0, *n*], the expected number of days of contagious exposure for an infected person is

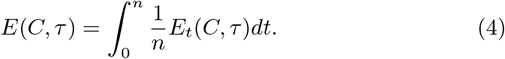

In the Appendix we show that Equations 3 and 4 imply the following proposition:

**Proposition 2.** *Given Assumptions 1 and 2, if n ≤ C − d*,

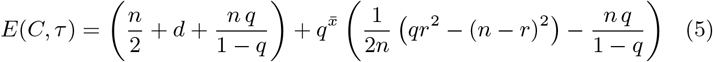

*where* 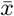 and *r are defined in Definition 2*.

We will use the formula that appears in Proposition 2 to calculate the exposure ratio and the estimated *R* values under some alternative testing regimes, under plausible assumptions about the parameters of infection.

## Self-isolation for some with symptoms

Even without testing, some individuals will choose to self-isolate once symptoms become evident. The number of days of contagious exposure for these individuals will include only those days when they are contagious before symptoms appear. With infection parameters *γ* = (*C, u, v, y*), the fraction (1 *− u*) of infected individuals will display some symptoms. Of these, the fraction *v* will self-isolate when symptoms appear. Thus the fraction of infected persons who self-isolate is *v*(1 *− u*) and the fraction who do not do so is 1 *− v*(1 *− u*). If infected individuals are contagious for a time period of *y* days before symptoms appear, then the expected amount of contagious exposure time for these individuals is *E*(*y, τ*).

It will be convenient to define the following functions that relate the parameters of infection and the testing regime to contagious exposure when some symptomatic individuals self-isolate.

**Definition 3** (Exposure functions). *Where γ* = (*C, u, v, y*) *and τ* = (*n, q, d*) *denote the parameters of exposure and of the testing regime, let E_nt_*(*γ*) *be the expected number of days of contagious exposure for an infected person with no testing, and let Ē*(*γ, τ*) *be the expected number of days of contagious exposure with testing regime τ*.

The expected number of exposure days for an infected individual is a weighted average of the number of exposure days for those who self-isolate and those who don’t. The fraction of infected individuals who self-isolate is (1 *− u*)*v* and the fraction who do not self-isolate is 1 *−* (1 *− u*)*v*. With infection parameters *γ* and testing regime *τ*, the expected number of days of exposure for an infected person who self-isolates will be *E*(*y, τ*) and for one who does not self-isolate will be *E*(*C, τ*). The expected number of days of contagious exposure for a randomly selected infected person is then:

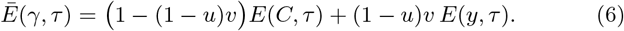

With infection parameters *γ* = (*C, u, v, y*), and with no testing, the expected number of days of exposure for an infected person will be

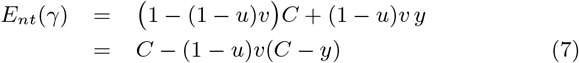

**Definition 4** (Exposure ratio function). *We define*

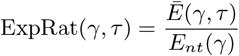

*to be the ratio of the expected amount of time that a contagious person is exposed to the public with testing regime τ to that if there is no testing*.

For any value of the reproduction number *R_nt_* that would apply without testing, we can use the expected exposure ratio to estimate the value of *R* that would apply given testing regime *τ*.

**Definition 5** (Estimated *R*-with-testing). *The function* 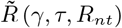 *is the estimated basic reproduction number R that results from testing regime τ, the vector of infection parameters is γ and where the R value without testing is R_nt_*.

Assuming that the expected number of people that one infects is proportional to the amount of time one is exposed to the population while contagious, we have the following simple relationship.

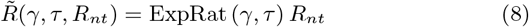

## Estimated exposure ratios and *R* values with testing

We used the contagion calculator found at https://steveli.shinyapps.io/ FrequencyAndAccuracyCalculator/ to explore the estimated exposure ratios and *R* values under alternative assumptions about the frequency, accuracy, and delay from test to isolation of testing regimes, given plausible infection parameters.

Tables 1–4 show estimated exposure ratios under various testing regimes. Throughout, we have assumed that in the absence of testing an infected person is contagious for *C* = 8 days, that those who display symptoms will do so after *y* = 3 days, that a fraction *u* = 0.4 of infected persons are asymptomatic, that *R* = 2.5, and that *v* = 0.3 of those who have symptoms will self-isolate without testing, four days after becoming contagious.

**Table 1:** Exposure ratio with delay *d* = 1.

| | $q$ | | | | |
| --- | --- | --- | --- | --- | --- |
|  | 0.1 | 0.2 | 0.3 | 0.4 | 0.5 |
| 1 day | 0.23 | 0.25 | 0.27 | 0.3 | 0.34 |
| 2 days | 0.31 | 0.34 | 0.39 | 0.43 | 0.49 |
| 3.5 days | 0.42 | 0.47 | 0.52 | 0.58 | 0.64 |
| 7 days | 0.63 | 0.67 | 0.71 | 0.75 | 0.79 |

**Table 2:**
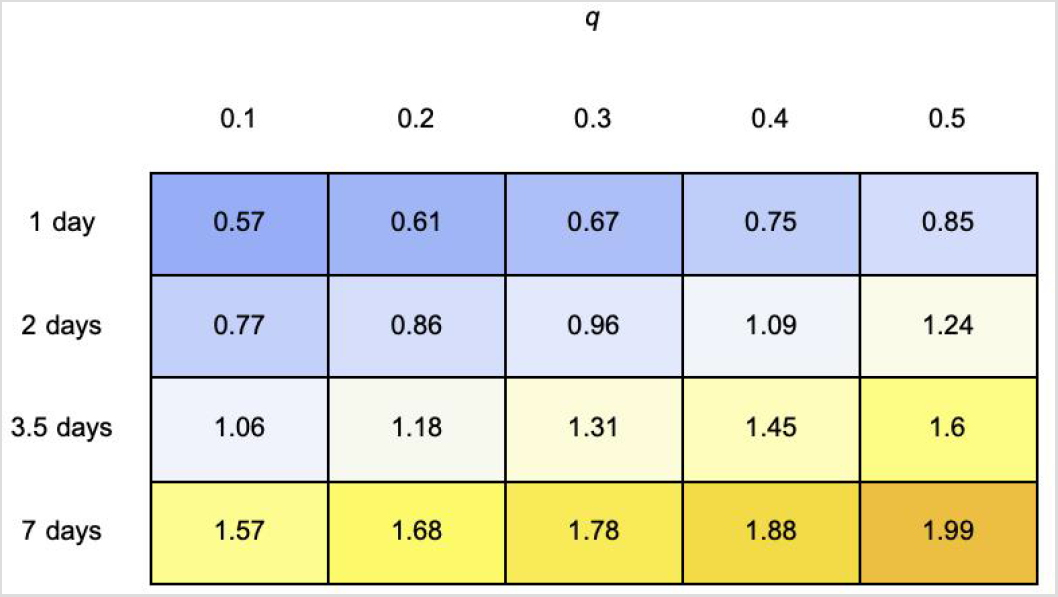
Estimated *R* if *d* = 1 and *R* is 2.5 without testing.

**Table 3:** Exposure ratio, report delay, and test frequency.

| | $d$ | | | | | |
| --- | --- | --- | --- | --- | --- | --- |
|  | 0 | 0.5 | 1 | 2 | 3 | 5 |
| 1 day | 0.13 | 0.27 | 0.41 | 0.53 | 0.61 | 0.57 |
| 2 days | 0.25 | 0.39 | 0.51 | 0.63 | 0.72 | 0.77 |
| 3.5 days | 0.4 | 0.52 | 0.63 | 0.73 | 0.81 | 0.89 |
| 7 days | 0.62 | 0.71 | 0.79 | 0.86 | 0.91 | 0.94 |

**Table 4:** Estimated *R*, report delay, and test frequency.

| | $d$ | | | | | |
| --- | --- | --- | --- | --- | --- | --- |
|  | 0 | 0.5 | 1 | 2 | 3 | 5 |
| 1 day | 0.33 | 0.67 | 1.01 | 1.32 | 1.54 | 1.43 |
| 2 days | 0.63 | 0.96 | 1.28 | 1.57 | 1.8 | 1.93 |
| 3.5 days | 1.01 | 1.31 | 1.58 | 1.82 | 2.04 | 2.21 |
| 7 days | 1.56 | 1.78 | 1.98 | 2.14 | 2.27 | 2.36 |

## Trade-offs between frequency and accuracy

Tables 1 and 2 illustrate the effects of test sensitivity on the exposure ratios and estimated effective reproductive number *R*, for various testing rates.

These tables indicate that if *R* = 2.5 in the absence of testing, a daily test with a one day delay could reduce *R* below unity even with a false negative rate of fifty percent. With a false negative rate of thirty percent, testing every second day would result in *R* < 1. With a false negative rate of ten percent, testing twice per week would get *R* very close to unity. Testing once per week would not bring *R* values close to 1.

## Trade-offs between delay and frequency

Tables 3 and 4 illustrate the effects of the delay between testing and isolation. These tables display the exposure ratio and estimated *R* for the stated test frequencies and delays, assuming a false negative rate of thirty percent.

The effect of reducing the delay between testing and isolation from two days to one day is similar to that of increasing test frequency from every second day to every day, or from twice a week to every second day. If the delay can be shortened from one day to half a day, exposure rates are further reduced by about twenty percent. The tables show that with a five-day delay, testing becomes almost worthless for reducing exposure and with a three day delay, even everyday testing falls short of reducing contagious exposure by fifty percent.

## Point-of-care testing

The US Food and Drug Administration has recently posted a template for commercial developers to help them develop simple COVID tests for which results could be determined on-site within minutes. In releasing this template, FDA Commissioner Stephen M. Hahn, M.D. said that^5^

> “We hope that with the innovation we’ve seen in test development, we could see tests that you could buy at a drug store, swab your nose or collect saliva, run the test, and receive results within minutes at home, once these tests become available.”

If such a test becomes available relatively cheaply, it would then be feasible to test very frequently with no significant delay between testing and isolation. Even if such a test had a relatively high probability of false negatives (low sensitivity), the ability to test frequently with quick response would permit frequent testing to drastically reduce the exposure ratio. Tables 5 and 6 show the exposure ratios and estimated *R* values as a function of test accuracy and frequency when test results are available immediately.

**Table 5:** Exposure ratio when test results are immediate.

| | $q$ | | | | |
| --- | --- | --- | --- | --- | --- |
|  | 0.1 | 0.2 | 0.3 | 0.4 | 0.5 |
| 1 day | 0.09 | 0.11 | 0.13 | 0.16 | 0.21 |
| 2 days | 0.17 | 0.21 | 0.25 | 0.31 | 0.37 |
| 3.5 days | 0.29 | 0.35 | 0.4 | 0.47 | 0.54 |
| 7 days | 0.52 | 0.57 | 0.62 | 0.68 | 0.73 |

**Table 6:**
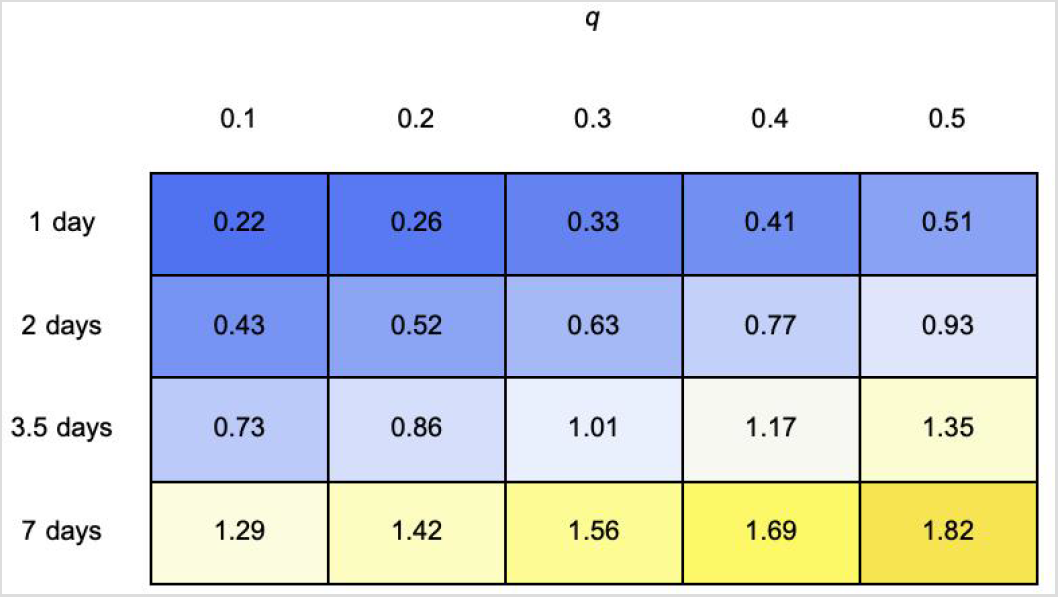
Estimated *R* when test results are immediate.

Table 5 shows that a test with very low sensitivity, if it can be administered every day with results appearing immediately, can drastically reduce the exposure rate. Indeed if those who test positive are quarantined immediately on taking a test administered daily, with sensitivity of 50%, the number of days of contagious exposure for infected persons is reduced by almost 80%. This compares with a reduction of about 60% that results from testing twice a week with a test with sensitivity of 90%.

## Discussion

As an increasing diversity of testing technologies receive emergency use authorization, communities are able to consider a broader range of possible testing strategies to contribute to COVID control. Any testing approach comes with tradeoffs: various tests differ in cost, convenience, sensitivity, inconclusive rate, and processing time.For example, saliva samples are easier to collect and less onerous to donate to collect than samples taken by nasopharyngeal swabs. Even if there were some reduction in sensitivity, these advantages could allow for more frequent collection and make saliva samples the preferred method. The use of pooled samples could substantially reduce the cost of testing, but might increase the fraction of false negative readings and possibly increase the delay between testing and reported results.^6^

To manage COVID within a community, it will be critical to assess how these tradeoffs play into optimal choice of testing strategies. While simulations can be useful in this endeavor, the sheer number of possible approaches will quickly overwhelm the ability to exhaustively explore all options using computationally intensive methods. The model and calculator we have presented here provide an alternative. Our analytic approximations can be used explore trade-offs between such variables as the frequency of testing, the sensitivity of testing, and the delay between testing and results.

Proactive testing is likely to be particularly important in college and university settings. Because students live in close proximity and engage in frequent interpersonal interactions, we would expect *R*_0_ on campuses to be high compared to nationwide averages. In addition, young adults appear more likely to experience subclinical disease [27] and thus less likely to self-isolate based on symptoms. Large clusters have already occurred in these university settings during summer sessions [28] and at schools attempting an in-person autumn semester [29]. In the absence of aggressive mitigation, colleges are likely to function as tinderboxes from which devastating epidemics emerge. Even if college-aged students are at lower risk for disease, on-campus clusters spread disease into more vulnerable populations in the surrounding communities [30].

Our results, and the results of other models[18, 31, 32], indicate that by proactively screening asymptomatic individuals and isolating those who test positive, universities should be able to substantially reduce the rate at which COVID spreads on campus. Yet the Centers for Disease Control chose to explicitly not recommend entry testing or ongoing testing of asymptomatic individuals on college campuses [28]. Taking this as absolution, many university administrators have failed to consider the possibility of frequently testing the entire student body and staff, on the grounds that doing is unnecessary, infeasible, and excessively costly costly^7^. A recent study examined the fall reopening plans of more than 50 universities as of August 7, 2020 [34]. The study reports that only about 27% of them plan to do some form of re-entry testing as students first arrive on campus and only about 20% plan to do some regular testing of all students.

Many of the exceptions are smaller East Coast institutions, a number of which are testing their students twice weekly in collaboration with the help of the Broad Institute [34]. The University of Illinois (UIUC) stands out among large public institutions in having implemented an in-house testing program that will administer saliva-based COVID-19 tests twice a week to each of its roughly 50,000 students and faculty.^8^ These tests, which have been developed in the university’s own laboratories have been found to be about as accurate as conventional PCR-based testing, and they offer approximately 5 hour turnaround between test administration and reported results.

Although many universities lack the resources and planning capability to apply regular testing and quarantine for their fall terms, cheaper and quicker tests appear to be on the horizon, and seem to be largely deterred by the lack of FDA approval [35]. Simple self-administered, paper-strip antigen tests have been developed by the Wyss Institute at Harvard and can be made available at a cost of $1-$5 per test. [35, 36]. As explained by Kotlikoff and Mina [35],

> You would simply spit into a tube of saline solution and insert a small piece of paper embedded with a strip of protein. If you are infected with enough of the virus, the strip will change color within 15 minutes.

These tests may not be as sensitive as standard RNA-based tests, but their cost and ease of testing would make it possible for them to be used by all community members every day, with results available immediately. Thus they are almost certain to be more effective in reducing the spread of the epidemic than more sensitive tests administered less frequently and with slower turnaround.

The model that we have developed here is analytically tractable because we have made certain simplifying assumptions about the nature of disease transmission. Perhaps foremost among these simplifications is the assumption that transmission rates are a step function: individuals who have COVID go from non-infectious to fully infectious instantaneously, and remain fully infectious until they are no longer able to transmit disease. Test sensitivity takes the same form over the course of infection. More sophisticated models could allow varying infectiousness and varying sensitivity over time, as in ref. [18]. In the absence of compelling data about the actual time-course of infectiousness and detectability, we have opted for the simple form analyzed here.

In our analysis we assume homogeneous susceptibility, transmissibility, and timing of the disease course across all individuals. Heterogeneity in disease parameters can have sizeable impacts on disease dynamics, but this is less of an problem than it might seem for what we are doing here. Our aim is not to explicitly model the dynamics of an outbreak, but rather to estimate the change in the frequency of transmission events. By doing so, we can compare the relative efficacy of alternative testing strategies and identify conditions under which testing will be sufficient to drop the effective reproductive number below unity. For this purpose, it is largely sufficient to work with mean parameter values. The major advantage to the approach we take here is that the analytic approximations enable more complex comparative static analysis and optimization efforts than would be feasible using time-consuming simulations. We hope that our findings will prove useful in this way to those involved in selecting testing procedures for colleges, workplaces, communities, and other groups.

## Data Availability

This is an analytic mathematical model. We have implemented a calculator for the formulas developed in the paper.

https://steveli.shinyapps.io/FrequencyAndAccuracyCalculator/

## Appendix

## Proof of Proposition 1

*Proof*.

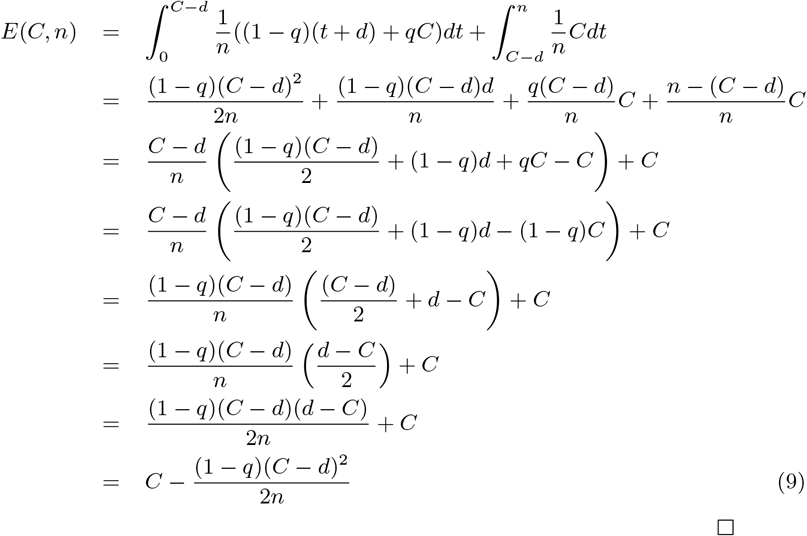

## Proof of Proposition 2

The following lemma will be useful for simplifying Equation 3.

**Lemma 1.** *Where* 0 *≤ q* < 1,

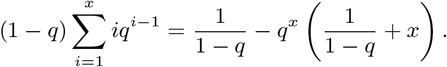

*Proof of Lemma 1*.

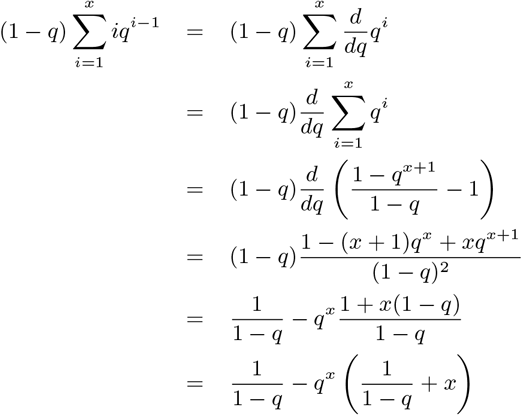

*Proof of Proposition 2*. From Equation 3 and Lemma 1, it follows that

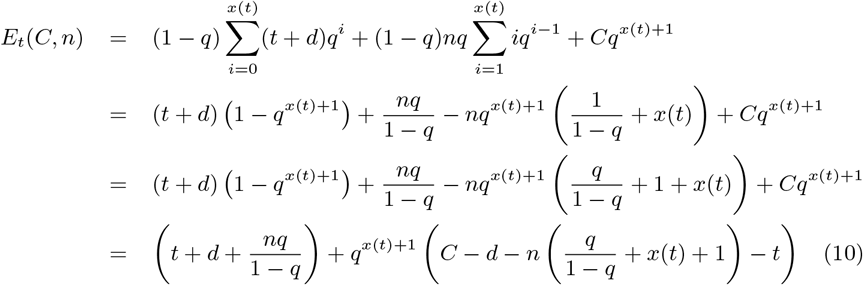

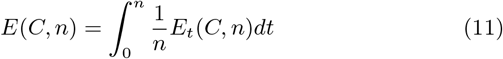

Equation 11 is more manageable than it first appears, because the function *x*(*t*) takes on only two values over its range. This is explained by the following lemmas.

**Lemma 2.** *For any A > n, if* 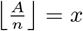 *and r* = *A − nx, then* 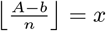 *if* 0 *≤ b ≤ r and* 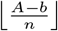 *if r < b ≤ r* + *n*.

Where 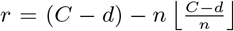, and *x ≤ r*, it follows from Lemma 2 that if *x ≤ r*, then 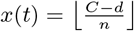 if *t ≤ r*, and 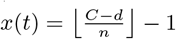. if *t > r*, Thus we have

**Lemma 3.** *Let* 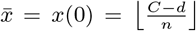. where 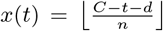 *and r* = 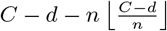, *we have* 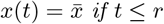 and 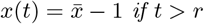.

Where 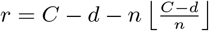, and 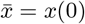, we have

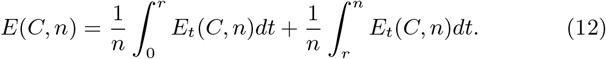

Then

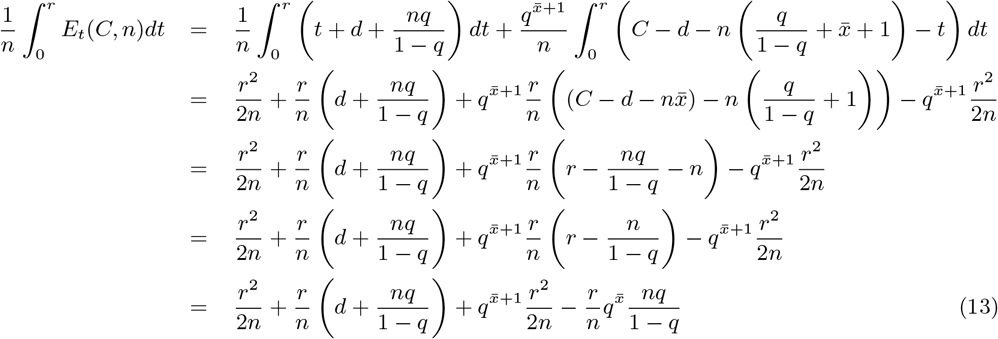

and

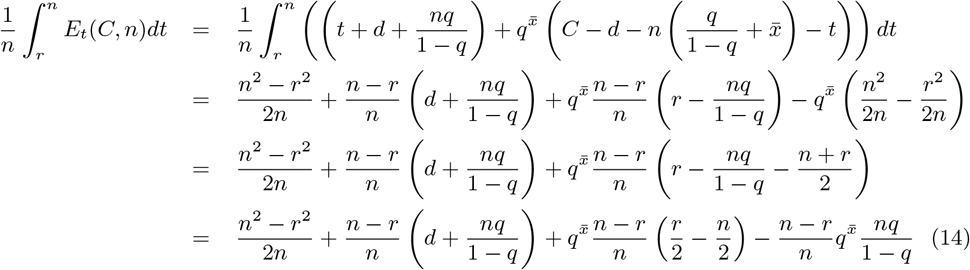

Equations 12, 13, and 14 imply that

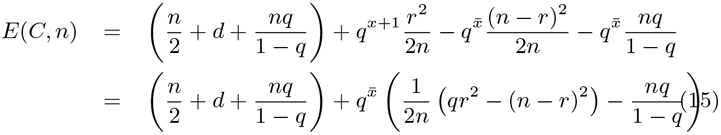

## Footnotes

1 https://www.cdc.gov/coronavirus/2019-ncov/hcp/duration-isolation.html

2 Kucirca *et al*. display a curve showing very high rates of false-negatives for presymptomatics, but these estimates seem to be meaningless, since they result from a curve-fitting exercise that uses only one pre-symptomatic observation.

3 According to the University of California at San Diego website, https://returntolearn.ucsd.edu/return-to-campus/testing-and-screening/index.html (accessed Sept 1, 2020), “The University is also planning to conduct periodic asymptomatic testing, most likely on a monthly basis.”

4 Even if testing occurs at fixed time intervals, the time at which infectiousness begins can reasonably be viewed as distributed continuously

5 Quoted on July 29, 2020 at https://www.fda.gov/news-events/press-announcements/ coronavirus-covid-19-update-fda-posts-new-template-home-and-over-counterdiagnostic-tests-use-non

6 In one basic approach to pooled sampling, half of the material from each subject is used in the pooled sample and half is withheld to be tested in case the pooled sample result is positive. If there is a positive result with the pooled sample, then the remainder of the material collected from each subject is tested. This retesting might add a half day or more to the time before the result are returned.

7 The president of the University of Michigan, Dr. Mark Schlissel maintains that for a large university such as Michigan, regular testing of students and staff is simply impossible. According to Dr. Schlissel, “This notion that a university at the scale of Michigan can test everybody a couple of times a week or every day, right now, that’s science fiction.” [33]

8 Current statistics on tests administered and the number of positive results are found at the UIUC dashboard https://splunk-public.machinedata.illinois.edu:8000/en-US/app/uofi_shield_public_APP/home.

